# The Gendered Implications of Energy Gaps in Health Care: A Comparative Analysis of Haiti, Senegal, and the Democratic Republic of Congo

**DOI:** 10.1101/2021.06.28.21259651

**Authors:** Vivek Shastry, Sophie M. Morse

## Abstract

The World Health Organization recently articulated a number of challenges faced by health systems due to unreliable access to energy services. Reliable energy availability at rural health facilities is understood to be an enabler of access to quality healthcare, owing to its potential impacts on medical services, health and safety, disease prevention and treatment, staff recruitment and retention, and administration and logistics. However, little is known empirically about the intersections of energy and healthcare, often due to the lack of availability of facility level data. Moreover, the gender implications of energy access (or lack thereof) for women as providers and seekers of primary healthcare have not been investigated. In this study, using a gender lens, we explore the linkages between energy and healthcare in three Francophone countries in the Caribbean and sub-Saharan Africa: Democratic Republic of the Congo (DRC), Haiti and Senegal. All three countries have faced serious challenges to the provision of quality health services, including infrastructure problems and specifically unreliable access to electricity. We use Demographic Health Survey data from all three countries to present detailed descriptions of the association between (a) the availability and reliability of electricity sources, and (b) availability of health services, equipment and medical personnel at different levels of the respective health systems. We find that the unavailability and unreliability of electricity is associated with lower availability of medical equipment and basic health services, especially among facilities at the primary care level in DRC and Haiti. Our findings highlight the opportunity to create more dependable and sustainable health systems by integrating decentralized clean energy technologies into health infrastructure, which can facilitate providers in female-dominated cadres such as nursing the ability to provide the care they are tasked with.

## 1. Introduction

Primary health care (PHC) addresses the majority of health needs throughout people’s lifetimes and is the cornerstone of a functional health system [1]. Ensuring access to high-quality primary care helps promote equity in health care, improve patient outcomes, and facilitate efficient use of resources [2]. One important element of PHC is the care provided to mothers, infants, and children. Although progress has been made in maternal and children health (MCH) care, in 2017 alone, 295,000 women died worldwide during and following pregnancy and childbirth [3]. Women in low and middle-income countries (LMICs) are at disproportionate risk. By providing access to quality preconception, prenatal, and inter-conception care for mothers and infants, the risk of morbidity and mortality in these populations can be minimized [4].

To ensure the availability of high-quality primary health care, there are various components that have to be in place, including access to energy. Recent work by the World Health Organization (WHO) shows that access to energy is associated with appropriate storage of vaccines and medicines, increased availability of health services, and enhanced health worker motivation among other benefits [5]. Nurses and community health workers (CHWs) are also crucial to the provision of PHC [6]. The impacts of unreliable access to energy relates to gender, both because of the importance of PHC for women and because of the care delivered by women in female-dominated professions at this level [7,8]. In this paper, through a gender lens, we investigate the relationship between access to energy and service delivery and the health workforce in three LMICs: Haiti, Senegal, and the Democratic Republic of the Congo (DRC).

The WHO building blocks Health Systems Framework allows for the evaluation and comparison of high-quality health services. It is comprised of six fundamental system building blocks: 1) Service delivery, 2) Health workforce, 3) Health Information Systems, 4) Access to Essential Medicines, 5) Financing, and 6) Leadership/Governance [9]. In developing countries, one important and often overlooked part of the building blocks and of guaranteeing universal health care includes access to efficient and reliable energy in health centers and hospitals [10]. United Nations Sustainable Development Goal (SDG) 3 on Health aspires to achieve universal health coverage by 2030, while SDG 7 on Energy aspires to ensure universal access to energy. Although their connection is not often highlighted, achieving SDG 7 is fundamentally tied to achieving SDG 3, because reliable energy availability is an enabler of access to quality healthcare owing to its potential impacts on medical services, health and safety, disease prevention and treatment, staff recruitment and retention, and administration and logistics. In this paper, we focus specifically on two building blocks impacted by energy access in different ways: the health workforce and service delivery (including equipment availability).

Women are the primary drivers of health globally, given their important roles in the health workforce [6]. Gendered norms influence the types of roles men and women have in the health system, as well as what roles are feasible and acceptable [7]. Globally, women are highly concentrated in nursing, midwifery, and primary care [6]. In order to do their jobs, among other factors, they need reliable energy access. It is essential for the provision of quality healthcare to women and children and this is especially true at primary and secondary tiers of health systems. Without it, health facilities at the community level may not have medical technologies that are required for the delivery of essential health services, such as vaccinations and safe births [10].

There are multiple ways that health facilities can be connected to energy sources. The most common type of energy source, especially in urban and peri-urban areas, is grid connected electricity supply. However, even with a grid connection, it is common for facilities to face constant power outages and failures [10]. The most common alternatives are stand-alone diesel-powered generators, which are expensive and logistically hard to maintain especially in remote rural regions. Another increasingly popular alternative is the use of solar powered electricity, either as stand-alone systems or in the form of community mini-grids. In certain sub-Saharan contexts such as Uganda, about 15 percent of hospitals use grid and solar electricity in tandem [11]. A grid-based electricity supply together with an alternate electricity source that can be used as a backup can provide comparatively more reliable energy access to the facilities. Facilities that either rely only on the grid, or only on non-grid alternate sources stand a higher chance of facing regular interruptions to supply. The worst off would be those facilities that do not have any access to electricity at all that might rely on candles or small battery powered lights if they need to deliver essential care after dark. Universal access to efficient modern energy services is rarely studied as an important condition for universal health coverage [10]. Our study contributes to this gap in research by presenting trends in both energy and primary care as well as interactions between the two across three countries and an exploration of the gender dimensions of the problem.

### 1.1. Country contexts

For the comparative analysis, Haiti, Senegal, and the Democratic Republic of the Congo were selected. These three French-speaking countries were all colonized by European countries (Haiti and Senegal by France and the DRC by Belgium), and all have persistent issues with access to quality health care, especially at the primary care level. Some key characteristics for each of the three countries are summarized in Table 1.

**Table 1.** Select Country Characteristics for Haiti, Senegal, and the Democratic Republic of the Congo.

| Country | Indicator |  |  |  |  |
| --- | --- | --- | --- | --- | --- |
|  | Population (thousands) 2019 | GDP per capita 2020 | HDI (rank) 2018 | MMR 2017 | IMR (out of 1,000 live births) 2017 |
| **Haiti** | 11, 263.08 | \$868.3 | .503 (169/189) | 480 | 49 |
| **Senegal** | 16,296.36 | \$1,522.0 | .514 (166/189) | 315 | 32 |
| **The DRC** | 86,790.57 | \$561.8 | .459 (179/189) | 473 | 67 |

Sources: Data were obtained primarily from the World Bank, the World Health Organization, and UNICEF [9,12–16].

All three countries rank low on the Human Development Indicator (a measure of human development based on life expectancy), and have low per capita gross national incomes, expected years of schooling among children, and mean years of adult education [17]. For the HDI, the DRC’s score is .459 (ranking 179/189), Haiti’s score is .503 (ranking 169/189), while Senegal’s score is .514 (ranking 166/189) [12]. The DRC has the lowest GDP per capita of the three at $561.8, while Haiti’s is $868.3, and Senegal’s is the highest at $1,522 [13]. While these three countries are at different stages of their development, they share persistent health problems at the population level as well as challenges to providing primary health care.

### 1.2. Population, health issues

#### Haiti

Haiti faces numerous health problems. Even before the 2010 earthquake that devastated the country, it was a “hot spot” for diseases such as rabies, malaria, and filariasis, all of which had long been under control in the rest of the Americas [18]. Life expectancy remains low: 61 for men and 66 for women [19]. Haiti also has one of the highest maternal mortality rates (MMR) in the region at 480/100,000 live births (in 2017) [20,21]. As compared to the Dominican Republic (a country sharing the same island), the mortality rate among children under five is times higher [2]. Infant mortality is also still 49/1,000 live births, which is more than three times higher than the regional average of 14/1,000 live births [15].

#### Senegal

Senegal fares slightly better than Haiti, but still faces myriad health challenges. A number of preventable communicable, neonatal, and nutritional diseases such as neonatal disorders, diarrheal diseases, lower respiratory infections, and tuberculosis are still the greatest contributors to premature death (all ages) [22]. Life expectancy is 65 for males and 69 for females, and the MMR is 315/100,000 live births [23,24]. While the infant mortality rate of 32/1,000 live births and under five mortality rate of 44/1,000 live births are both lower than the averages for sub-Saharan Africa, there is still room for improvement through improved access to quality care [16].

#### The DRC

The DRC is lagging behind most countries in the world in terms of basic health indicators. Malaria is still the number one cause of mortality (all ages), and similar to Senegal, other preventable diseases such as lower respiratory infections, neonatal disorders, and tuberculosis are the other top causes of death [25]. Unsurprisingly, life expectancy is 60 for males and 64 for females. The DRC is also one of the top five contributors to maternal and child mortality in the world [26]. The MMR is 473/100,000 live births, and infant mortality is the highest of the three countries at 67/1,000 live births [21,27]. Under five mortality remains 81.9/1,000 live births, which is slightly higher than the sub-Saharan African average of 80/1,000 live births [1].

### 1.3. Health system overviews

In order to provide a context for comparative health system analyses, we summarize how each health system is structured. Below we provide an overview of each health system and its salient issues.

#### Haiti

The Haitian health system is organized into three levels: 1) primary care, which is provided at around 800 health centers and 45 community reference hospitals; 2) secondary care, which is provided at 10 departmental hospitals; and 3) tertiary care, which is provided three specialized centers and give university hospitals [28]. Prior to the 2010 earthquake, Haiti already had one of the weakest health systems in the western hemisphere [18]. The earthquake devastated an already fragile system. Although there has been major investment from external actors such as USAID and the Centers for Disease Control and Prevention (CDC) to fix the problems caused and exacerbated by the earthquake, health system issues persist [29,30]. The public spending on healthcare remains some of the lowest in the world [2]. Moreover, primary health care including antenatal and postnatal care and vaccinations are not used consistently by Haitians. For example, the routine coverage for the measles-rubella vaccination was only 58 percent [2,18].

#### Senegal

In Senegal, there are three “pyramid” levels that form the structure of the health system: central, regional, and peripheral. The central level comprises the Ministry of Health and health branches, and related services [31]. The regional level refers to the local health system, and the peripheral is made up of health districts with at least one health center and a network of smaller health facilities [31]. There are four levels of facilities: 1) Health Posts (Poste de Santé); 2) District Health Centers (Centre de Santé); 3) Regional Hospitals (Hospital/Clinic Provincale) (specialized health centers); and national level hospitals [31,32]. Although there have been great improvements in health care coverage in Senegal and significant decreases in malaria cases and deaths, evidence shows that the quality of primary care varies greatly [33].

#### The DRC

Similar to Haiti and Senegal, the DRC’s health system is organized into three hierarchical levels: the central level, the intermediate level, and the implementation level [34]. The central level’s role is normative, while the intermediate level is managed by provincial health departments and plays a technical and logistical support role. The implementation level consists of 516 health districts, with a network and health centers and district hospitals managed by district teams [34]. The health system in the DRC was weakened by more than a decade of conflict and due to this fragile context, the state is unable to widely deliver quality care to the population [35,36]. Similar to Haiti, government expenditure on health per capita is one of the lowest in the world [37]. Often, drugs and personnel are unavailable and given the low investment and poor financial management, providers rely on unofficial payments by users as well as high user fees [35]. As a result, in the DRC, there are still excessive fee-for-service payments for primary health services [36].

## 2. Materials and methods

In the context of these fragile primary health systems, little is known about the intersections of energy and healthcare due to the lack of consistent availability of facility level data for all countries. We utilize USAID’s Service Provision Assessment (SPA) dataset, which is a subset of the Demographic Health Survey (DHS) that collects detailed health system [38]. This allows us to study the intersection of energy and health services. To focus the analysis for this paper, we restricted the geographic scope to countries in the African Subcontinent and the Caribbean region. We further selected only those countries for which data were available in the past five years (2015-2020) so that our findings reflect the current situation. Only three countries met these criteria: The DRC (2017-18), Haiti (2017-18), and Senegal (2018). We utilize these data to study the variation in the availability of health services among facilities with different levels of electricity access in the three selected countries.

We present the summaries from the DHS SPA datasets in three stages. First, we focus on the availability of basic health services across different tiers of the health system in each country. Specifically, we compare the availability of services that are essential to deliver safe mother and child care, such as vaccinations, normal/vaginal deliveries, lab services and cesarean sections.^1^ Second, we focus on the availability of electricity at health facilities. Here we also summarize the types of electricity sources and the reliability of power supply at the facilities. Finally, we focus on the primary (or the lowest) level health facilities in each country to describe the variation of average health service availability across facilities with different types of electricity sources. In addition to the variation of health service availability, we also describe the variation of medical equipment and workforce availability across these facilities. For the medical equipment, we focus on the basic equipment necessary for safe childbirth, such as delivery lights, incubators, ultrasound and electric autoclaves. For the workforce, we compare the availability of doctors (full time and part time), nurses (professional and degree) and midwives (professional and degree). For each category, we compute and plot the mean along with a 95 percent confidence interval, as shown in all figures in section 3.3. When the confidence intervals don’t overlap, we interpret the means to be statistically significantly different [39].

In this study, we present descriptive breakdowns of variation in health services and energy access and specifically focus on the availability of health services, basic medical equipment and workforce at primary care facilities with different levels of energy access. We acknowledge two substantial limitations of this approach. First, there are many other factors that determine the availability of health services, including and not limited to the availability of adequate funding, remoteness of the facilities, and other infrastructure considerations such as stable buildings, running water and resident quarters for medical staff. Some of these could be confounding factors that are associated with both the variable of interests here - the availability of health services and the availability of reliable electricity access. In this study, due to limitations of the data, we do not control for these variables and we do not investigate the causal relationship between the variables. By focusing on the descriptive analysis, we aim to describe the problem and lay the groundwork for future studies that can estimate the impact of poor electricity access on health services after controlling for other covariates. Another recent study has developed a methodology to estimate these impacts [8]. Second, the mere availability of a particular health service at a facility is insufficient information to draw inferences about the delivery and utilization of that service at that facility. The availability of staff and resources to deliver that service, the quality of service delivery and the extent to which households are willing to utilize that service are all relevant factors that determine the successful delivery of a health service. Since the survey does not capture the quantity, quality or utilization rate of these services, we are not able to offer granular insight beyond just the availability of services.

## 3. Results

### 3.1. Status of health service availability

In this section, we present the basic health services that were reported to be available in facilities in each respective country. For the context of this study, basic health services include vaccination, normal delivery, caesarean section (C-section), and lab services. We present the percentages of the total numbers of each type of health facility in which the respective health service was reported to be available.

#### Haiti

Inadequate availability of basic health services in Haiti is evident from the data in Table 2. One in five dispensaries and community health centers in Haiti indicated they were not offering vaccinations. Only a third of these centers had laboratory services available, and only a fifth of the centers were conducting normal deliveries. Even among health centers with beds, a third of these facilities were not conducting normal deliveries. Almost none of the facilities at the primary care level had the ability to conduct C-sections, showing that many women, especially those with complications, have to travel from remote areas to the nearest community reference hospitals to deliver their babies. The availability of basic health services at the secondary and tertiary levels, including community reference hospitals, departmental hospitals and university hospitals, was much better relative to the primary care level.

**Table 2:** Percentage of health facilities that reported the availability of basic health services.

| Country | Health Center | Vaccination | Normal Delivery | C-Section | Lab Services |
| --- | --- | --- | --- | --- | --- |
| **Haiti** | Dispensary or Community Health Center (n = 352) | 80% | 19% | 0% | 31% |
|  | Health Center without bed (n = 361) | 59% | 21% | 1% | 73% |
|  | Health Center with bed (n = 163) | 68% | 64% | 7% | 80% |
|  | Community Reference Hospital (n = 46) | 85% | 94% | 70% | 98% |
|  | Departmental Hospital (n = 8) | 100% | 100% | 88% | 100% |
|  | University Hospital (n = 6) | 100% | 100% | 100% | 100% |
|  | Other Hospital (n = 71) | 36% | 70% | 60% | 90% |
| **Senegal** | Health Post (n = 73) | 77% | 47% | 0% | 0% |
|  | Health Center (n = 32) | 71% | 63% | 8% | 99% |
|  | Clinic (n = 119) | 88% | 82% | 0% | 92% |
|  | Hospital (n = 33) | 38% | 78% | 65% | 78% |
| **The DRC** | Health Center (n = 540) | 95% | 95% | 11% | 81% |
|  | General Referral Hospital (n = 483) | 41% | 96% | 96% | 96% |
|  | Referral Health Center (n = 214) | 94% | 98% | 80% | 96% |
|  | Hospital or Hospital Center or Clinic (n = 138) | 61% | 93% | 88% | 99% |
|  | Territorial or Provincial Hospital (n = 5) | 80% | 100% | 100% | 100% |

#### Senegal

Health service availability at the primary care level in Senegal is observed to be similar to that of Haiti. A third of the health posts did not offer vaccinations, and only half of them were conducting normal deliveries. while none offered laboratory services. The status of secondary level health centers and clinics was similar with respect to vaccinations with 71% of health centers and 88% of clinics providing them. Most of these centers offer lab services, whereas a third of the health centers and a fifth of the clinics were still not conducting normal deliveries. Almost none of the health facilities at the primary and secondary level conducted C-sections (less than 10%), and even at the tertiary level hospitals, a third of them were not offering this service. This demonstrates that safe childbirth is likely very difficult and stressful for pregnant women with even minor complications.

#### The DRC

According to the survey, basic health services were available at most of the health facilities (Table 2). The availability of vaccination was high in primary level health centers (95 percent), and considerably lower in the referral hospitals (41 percent) and hospital centers (61 percent). On the other hand, the availability of basic laboratory services was slightly lower among health centers (81 percent). Normal deliveries were reported to be widely available across all health facilities and even C-section was reported to be available at most facilities except at the primary level health centers.

### 3.2. Status of electricity supply availability

In this section, we present the status of electricity supply availability for each type of facility in the three countries. Figure 1 shows the main sources of electricity by facility type and Figures 2 and 3 present the alternate sources of supply and interruptions to power supply respectively. Overall, we observe that while most of the tertiary level facilities have grid-based electricity supply available along with one or more alternate sources as backup, most of the primary level facilities across the three settings rely substantially on only alternate (non-grid) sources of electricity.

**Figure 1:**
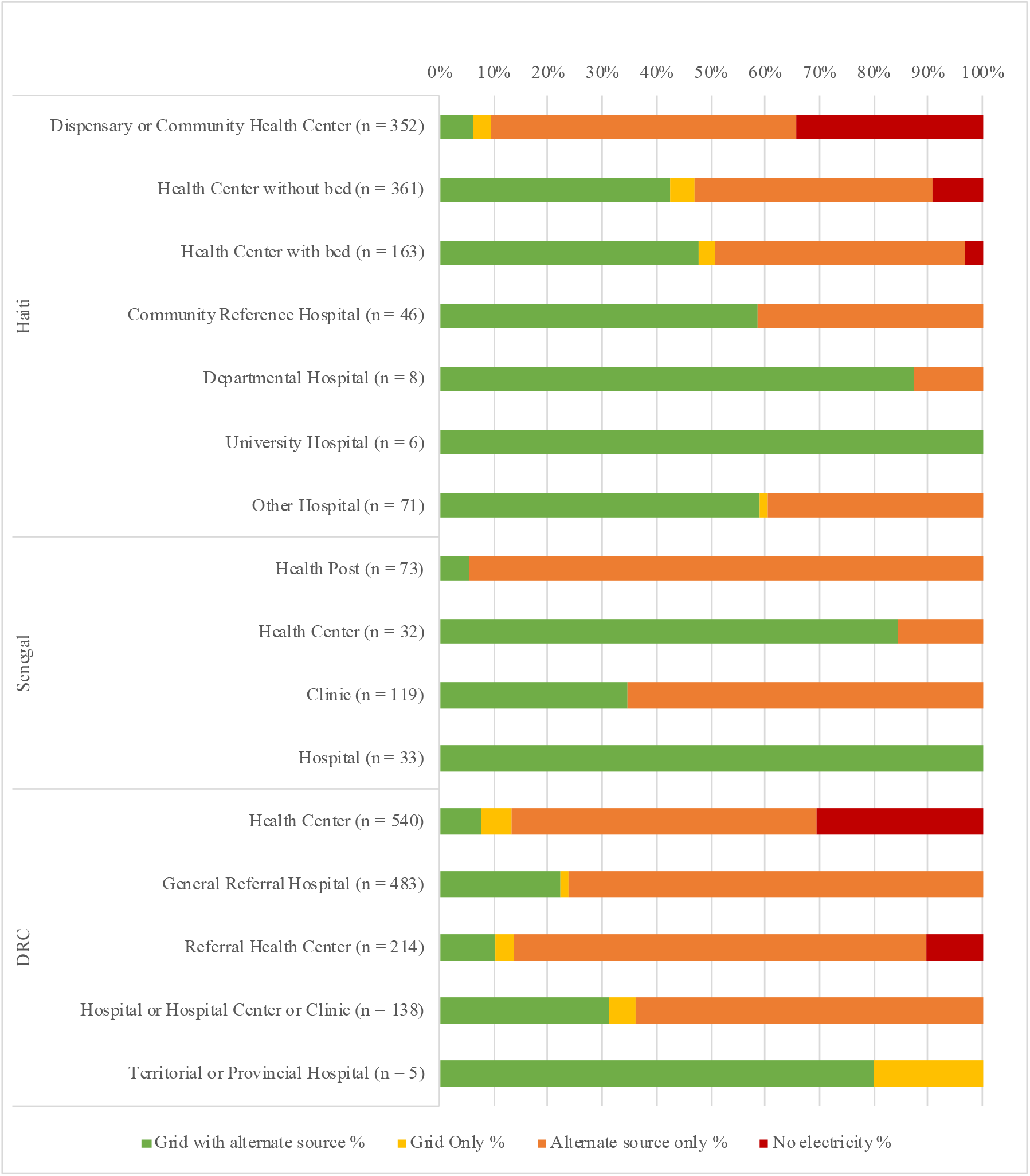
Percentage of Health Centers according to Sources of Power Supply.

**Figure 2:**
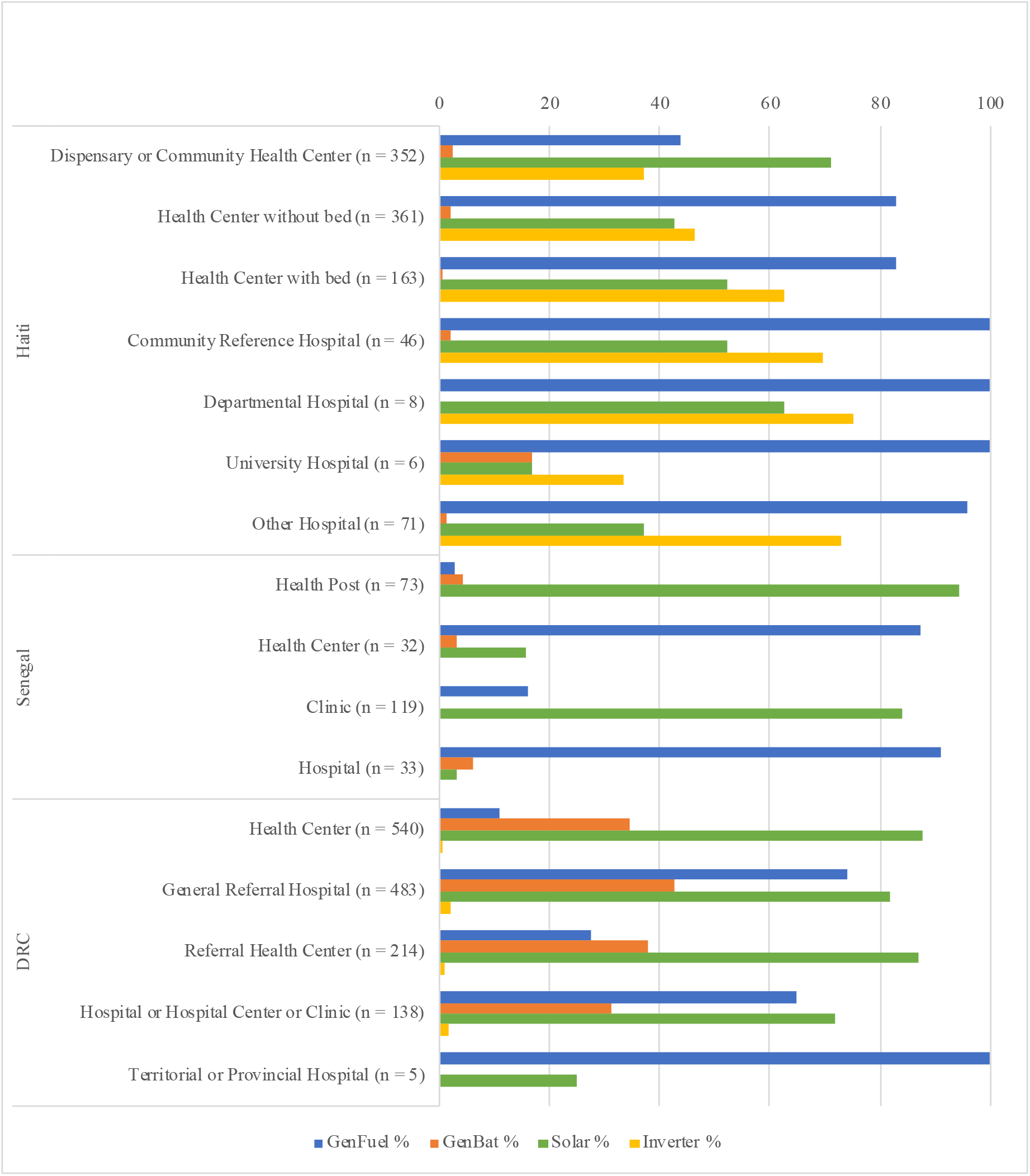
Percentage of Health Centers having Alternate Sources of Power Supply.

**Figure 3:**
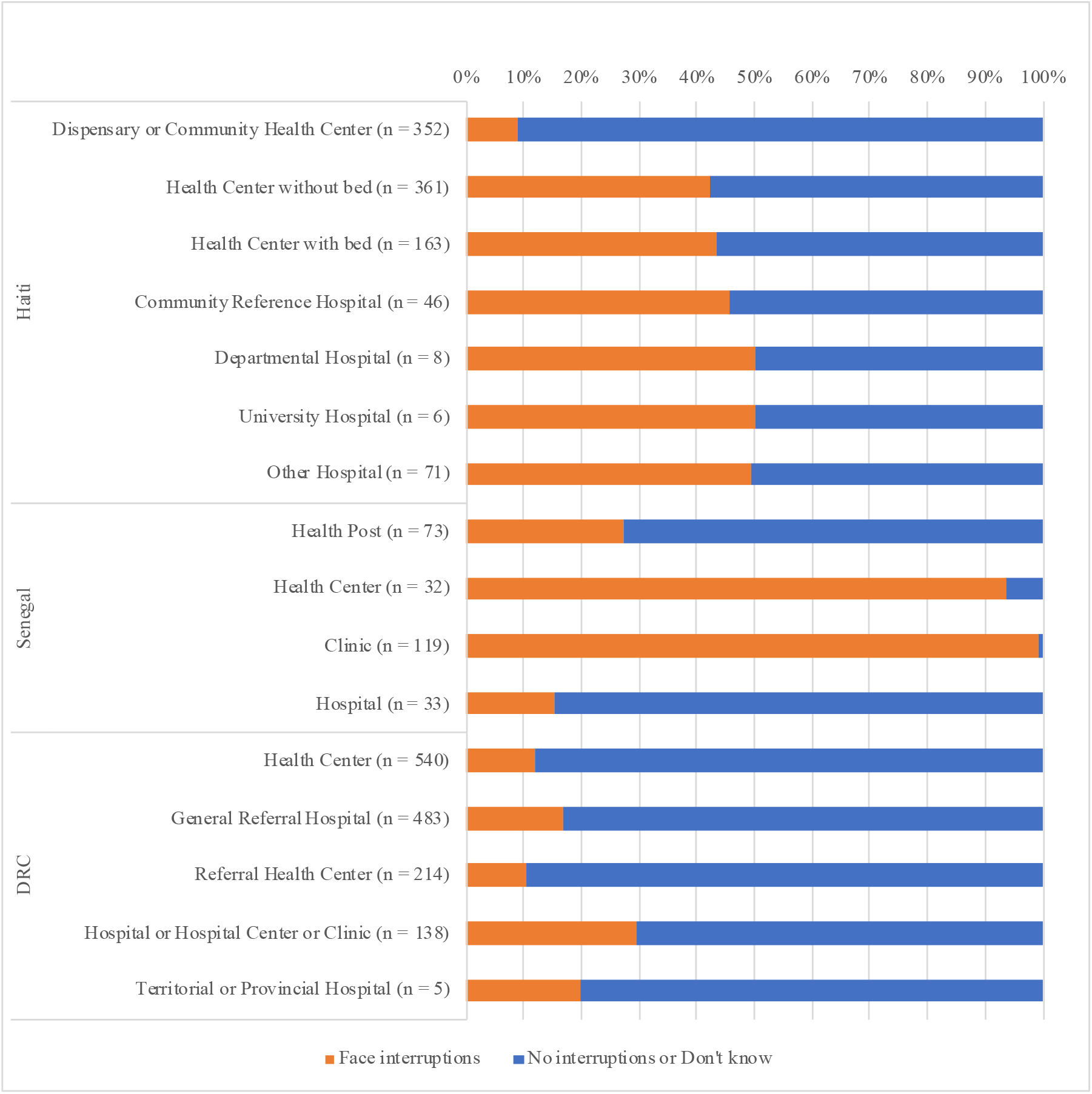
Percentage of Health Centers facing interruptions in Power Supply.

#### Haiti

In Haiti, a third of the primary level dispensaries or community health centers did not have any access to electricity at the time of survey. Most of those that did have electricity relied primarily on non-grid alternate sources. One-tenth of the health centers without beds also did not have electricity access. Except at the primary level, more than half of the facilities at all other levels had access to grid supply, together with an alternate source of electricity. Fuel-based generators were the main source of backup supply, but more than half of the facilities also used inverters and solar systems. Almost two-thirds of the primary level dispensaries had access to solar based electricity. Half of all facilities except at the secondary and tertiary levels reported regular interruptions in supply.

#### Senegal

The availability of grid supply for the health posts at the primary care level was extremely poor, with 95 percent of the facilities having to rely exclusively on alternate sources of electricity. While most of the health centers and hospitals did have access to grid supply, only a third of the clinics had grid access. The use of solar as an alternate energy source was prominent in health posts and clinics, whereas fuel-based generators were more common in health centers and hospitals. On a positive note, all of the facilities surveyed in Senegal had at least one source of electricity. However, a third of all the health posts and almost all the health centers and clinics said they faced regular interruptions in supply.

#### The DRC

A third of the primary level health centers and ten percent of the referral health centers reported not having any electricity at all at the time of the survey. Grid-based electricity was prominent only in tertiary level hospitals, while less than a third of the remaining facilities had access to the grid. Over half of the primary and secondary level facilities were powered only through alternate sources. Almost four out of five facilities at the primary and secondary level reported having solar technologies, making it the most prominent alternate electricity source in the DRC. This was followed closely by fuel generators, more so at secondary and tertiary facilities. The use of inverters was minimal in the DRC. In general, most facilities reported a relatively low level of interruptions in electricity supply: only ten to thirty percent of the facilities said they faced regular interruptions in supply. Since the survey doesn’t distinguish between “Didn’t know” and “No interruptions,” we are not able to infer if the remaining facilities that were surveyed did not have any interruptions, or just did not know about it.

### 3.3. Interactions between electricity and health services

We observe in section 3.2 that across settings the availability of electricity is poorer in the health facilities at the primary care level as compared to the secondary and tertiary level clinics and hospitals. In this section, we investigate how the health service availability differs for health facilities that have different levels of electricity access specifically at the lowest level primary care centers. We therefore focus on community health centers in Haiti, dispensaries or and Health posts in Senegal, and health centers in the DRC.

#### Haiti

Among the dispensaries in Haiti, we observe significant variation in the availability of lab services (Figure 4). While 75 percent of the dispensaries with grid access and alternate sources offered lab services, only around 40 percent of dispensaries relying only on alternate sources and less than 15 percent of those without electricity offered lab services. The availability of normal deliveries is also observed to be higher in dispensaries relying only on alternate sources compared to those without electricity and even those with grid access. Almost none of the dispensaries regardless of the electricity status had access to any of the four pieces of basic medical equipment, not even a delivery light or an autoclave (Figure 5). Even among dispensaries with access to grid and alternate sources, less than 25 percent of them had access to an electric autoclave. In terms of workforce, we notice that dispensaries without electricity did not have a full-time doctor at all, whereas dispensaries with grid access and alternate sources had at least one full time doctor available on average (Figure 6). Similarly, the dispensaries with better electricity access had four times as many degree midwives on average.

**Figure 4:**
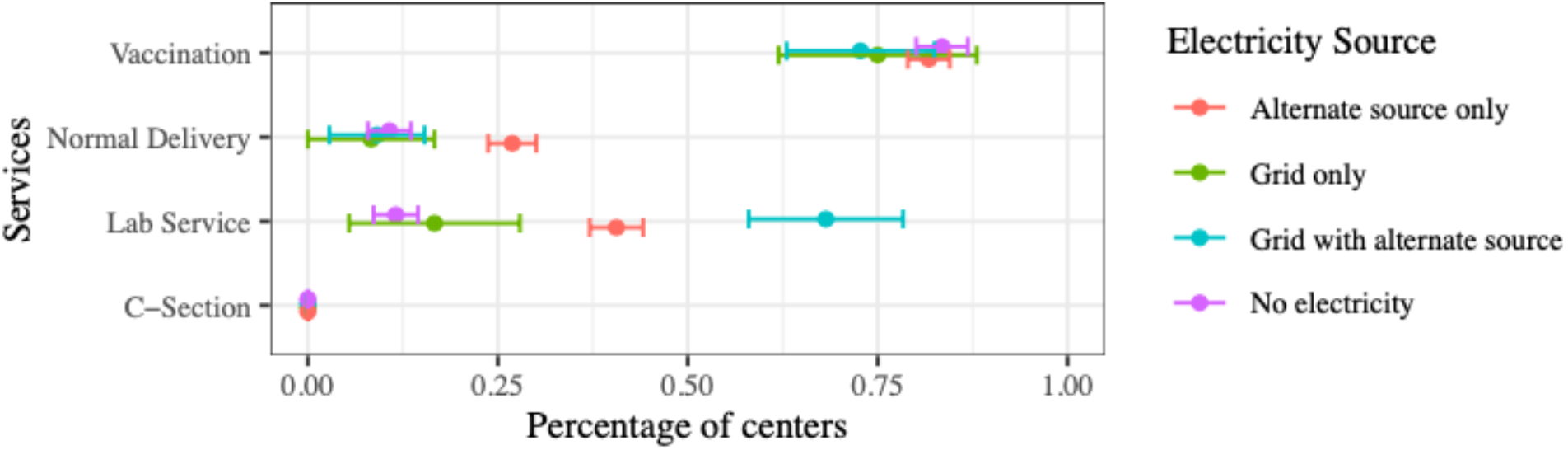
Percentage of primary level dispensaries or community health centers in Haiti with availability of basic health services, categorized by type of electricity source (N=352) Note: Error bars represent a 95 percent confidence interval for the mean.

**Figure 5:**
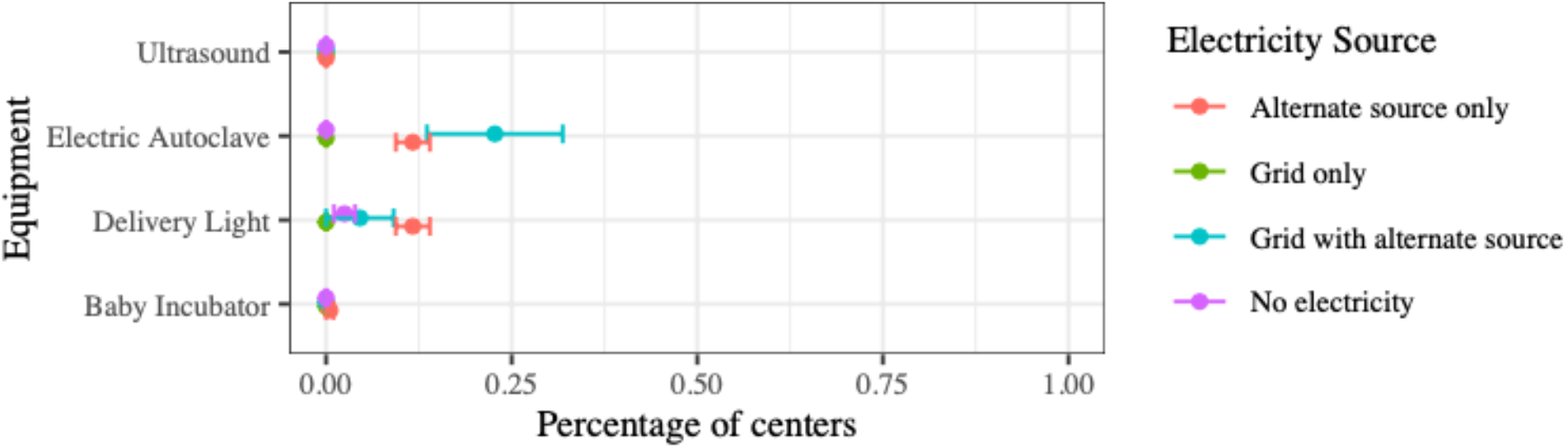
Percentage of primary level dispensaries or community health centers in Haiti with availability of basic medical equipment, categorized by type of electricity source (N=352) Note: Error bars represent a 95 percent confidence interval for the mean.

**Figure 6:**
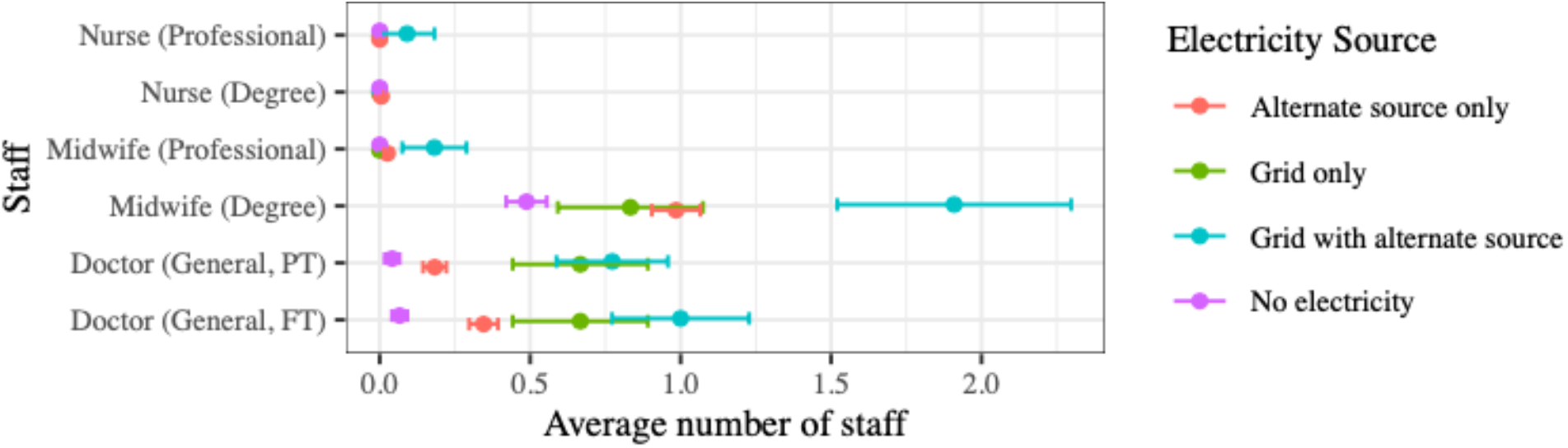
Average number of health workforce present at primary level dispensaries or community health centers in Haiti, categorized by type of electricity source (N=352) Note: Error bars represent a 95 percent confidence interval for the mean.

#### Senegal

As noted in section 3.1, all health facilities in Senegal do report having some form of electricity access. In Figures 7 and 8, we see that among the health posts at the primary care level, there are no statistically significant differences in health services or equipment availability. Overall, these health posts were not providing any lab services, and only 25 percent of the facilities reported having access to a delivery light. This indicates that most health posts are not equipped to deliver comprehensive mother and childcare, especially deliveries. Workforce data for health posts were not available in the Senegal survey.

**Figure 7:**
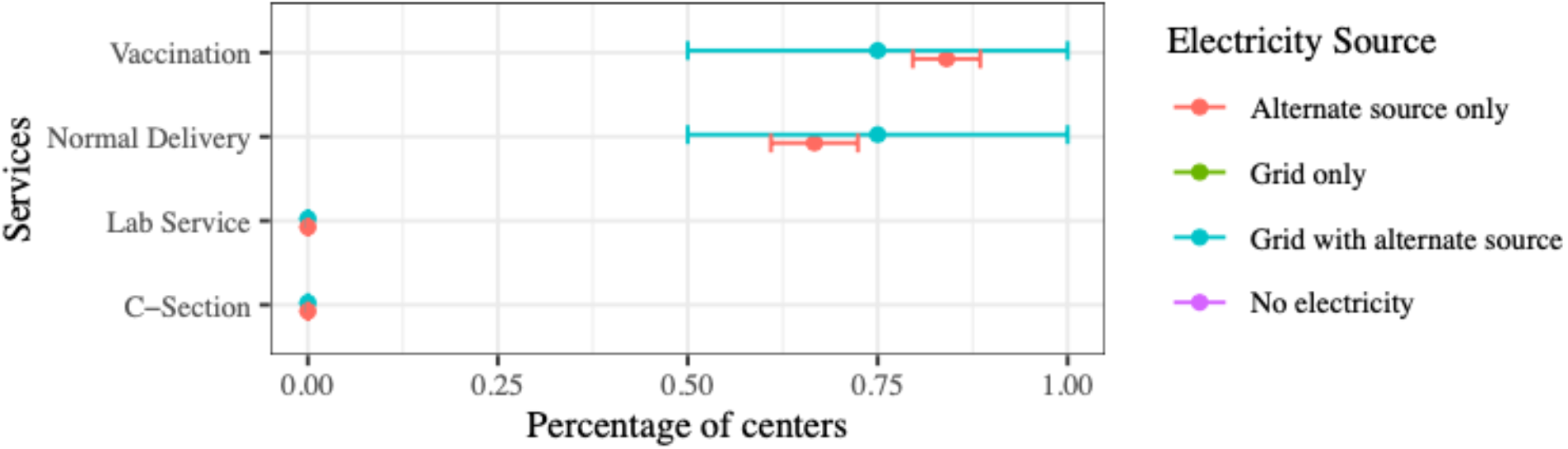
Percentage of primary level health posts in Senegal with availability of basic health services, categorized by type of electricity source (N=73) Note: Error bars represent a 95 percent confidence interval for the mean.

**Figure 8:**
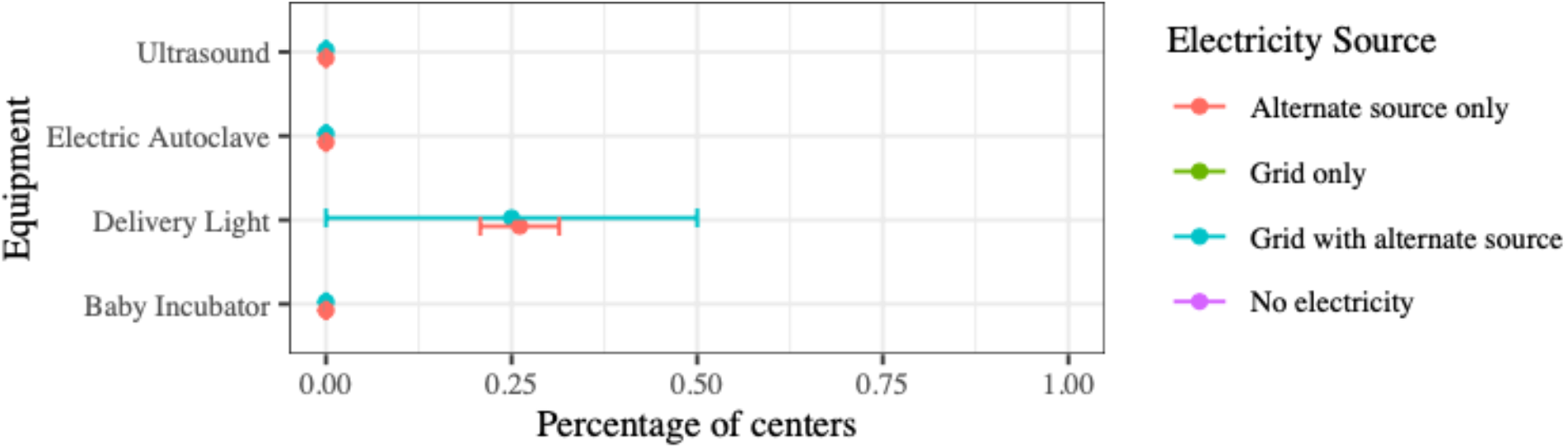
Percentage of primary level health posts in Senegal with availability of basic medical equipment, categorized by type of electricity source (N=73) Note: Error bars represent a 95 percent confidence interval for the mean.

#### The DRC

The availability of vaccination and normal deliveries were roughly the same for all types of electricity access in the DRC (Figure 9). However, only 75 percent of the health centers without electricity offered lab services whereas almost all of the health centers with grid access and alternate sources did. The difference is also noticeable for availability of cesarean sections— one in three health centers with grid access and alternate sources offered C-sections, whereas almost none of centers without electricity access did. Seventy five percent of the health centers with grid access and alternate sources had delivery lights available, whereas only 50 percent of centers without electricity access did (Figure 10). Similarly, one in three health centers with grid access and alternate sources had access to ultrasound devices whereas almost none of centers without electricity access did. We notice a variation in workforce availability as well. While health centers with grid access and alternate sources had 2 full time doctors and 6 degree nurses on an average, centers without electricity access had no full time doctors and only 2 degree nurses on an average (Figure 11). The situation at health centers powered only by non-grid alternate sources was very similar to health centers that had no electricity at all.

**Figure 9:**
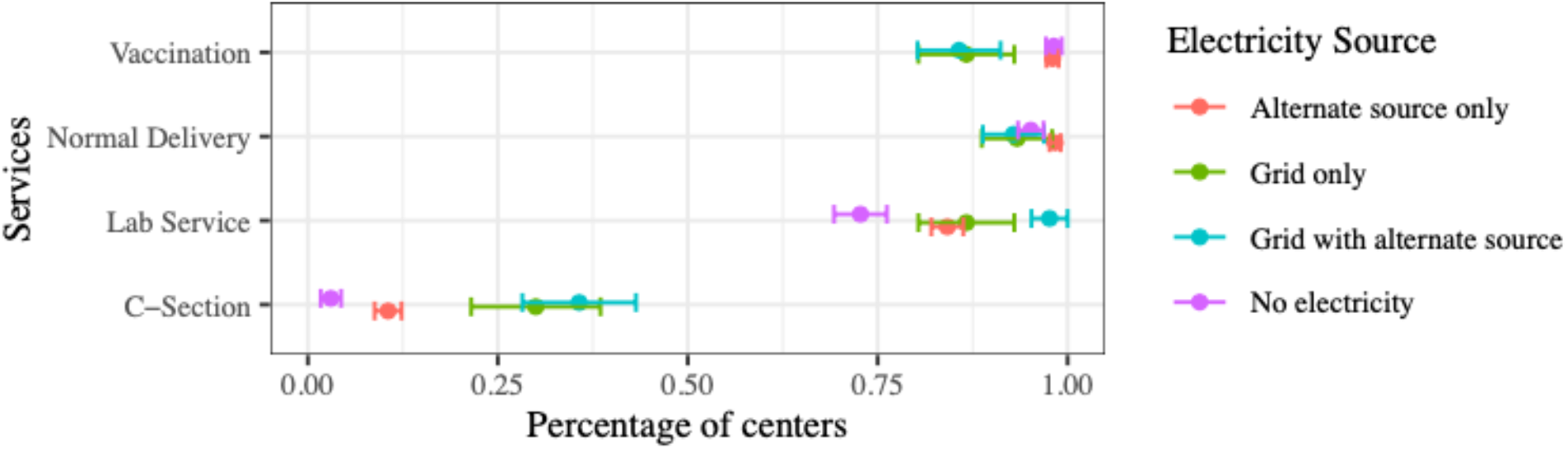
Percentage of primary level health centers in the DRC with availability of basic health services, categorized by type of electricity source (N=540) Note: Error bars represent a 95 percent confidence interval for the mean

**Figure 10:**
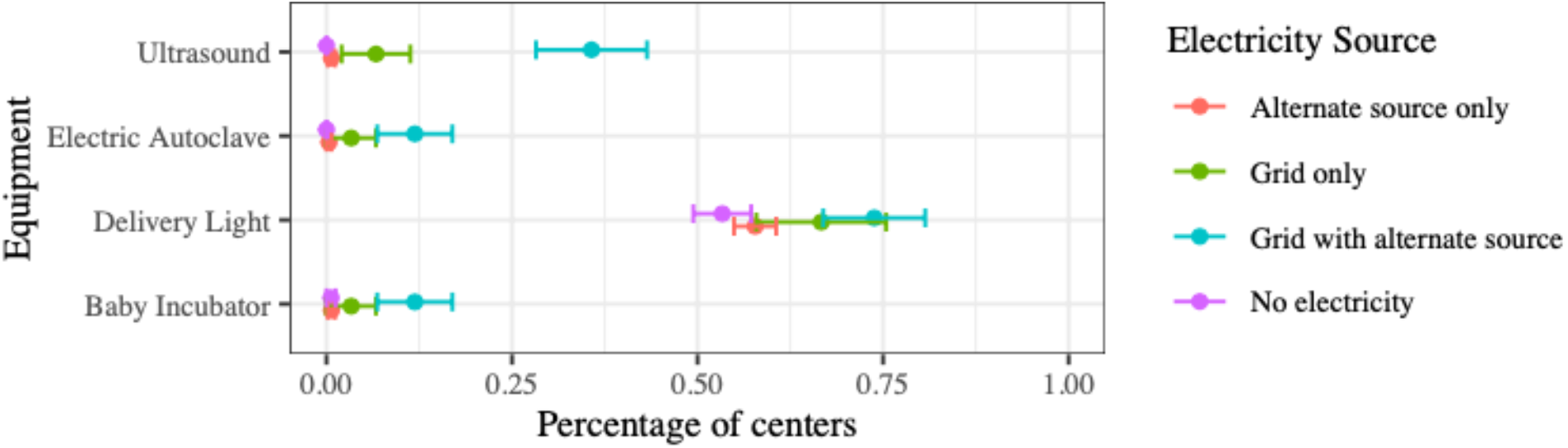
Percentage of primary level health centers in the DRC with availability of basic medical equipment, categorized by type of electricity source (N=540) Note: Error bars represent a 95 percent confidence interval for the mean

**Figure 11:**
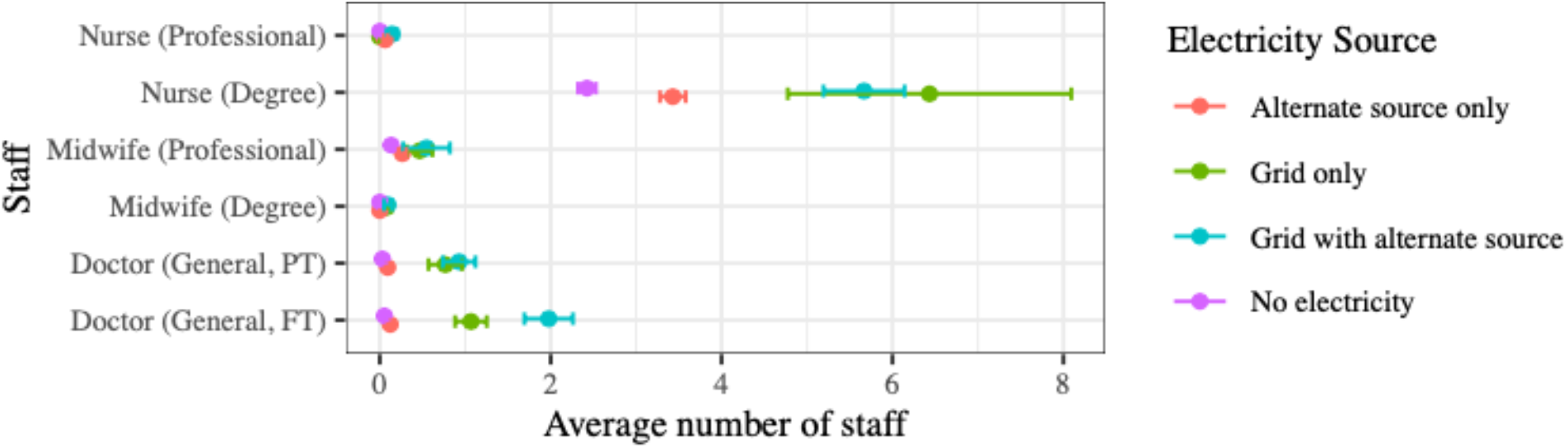
Average number of health workforce present at the primary level health centers in the DRC, categorized by type of electricity source (N=540) Note: Error bars represent a 95 percent confidence interval for the mean.

## 4. Discussion

Across the three countries included in this study, we observe disparities with various health system building blocks including service delivery and the health workforce, as well as gaps in energy availability and reliability. We find substantial variation and reduced availability of vaccinations, lab services, normal deliveries, and C-sections at the lower tiers of the health system. This is consistent with work on service readiness across multiple LMICs (including Haiti and Senegal), which finds that across settings the mean value on a service readiness index was 52 percent for health/centers and clinics as opposed to 77 percent for hospitals [40]. Service availability in Haiti and Senegal was generally lower in comparison to the DRC, which is somewhat surprising based on our review of health system challenges in that setting. At the primary level we observe relatively lower availability of some types of medical equipment and the same is true for health workers in the DRC and Haiti, which can translate to poorer quality of care.

Energy access is also less reliable in lower tiers of all three health systems. Most of the primary level facilities across all settings rely substantially on only alternate (non-grid) sources of electricity, with Haiti and the DRC lacking any electricity at one third of their primary level health centers. In Senegal, almost all health posts had some type of electricity access and 95 percent of facilities relied on alternate sources. This is consistent with research from WHO highlighting that hospitals tend to be located closer to electricity grid connections, whereas health centers at the primary level tend to rely more on generators or alternative sources such as solar power [10]. Prior research from Haiti and Senegal also that shows that hospitals have better service readiness around basic amenities such as water and electricity when compared to clinics and health centers [40].

The relationship between electricity access, service availability and the health workforce is more nuanced and varies by country. In Senegal there were low levels of lab services irrespective of electricity source whereas in both Haiti and the DRC the availability of lab services was higher in settings with some sort of grid access. Such a difference was not observed for vaccinations or normal deliveries. However, we did observe that the availability of basic medical equipment to conduct deliveries, such as autoclaves, delivery lights and ultrasounds was lower among primary health facilities without electricity access in both Haiti and the DRC. For the health workforce, in Haiti and the DRC health facilities with grid sources of electricity have higher availability of midwives and doctors and in the DRC the same is also true of nurses. This has been reflected in previous studies showing shortages of trained healthcare workers: for every 3,000 Haitians, there is only one medical doctor and one nurse and physicians are largely concentrated in Port-au-Prince, the capital city [2,41]. Community Health Workers (CHWs) are being used to help fill this health worker crisis [42]. In the DRC, there have also been challenges with maintaining a well-supported and financed public workforce [43]. Although we did not have data on staff availability in Senegal, similar to Haiti, health care workers are concentrated in the capital of Dakar and there is only one trained physician for every 100,000 people [33,44].

There are gendered implications of differential energy access in health services with regards to the health workforce. It is well-documented that globally, women are concentrated in primary care, nursing, and midwifery [6]. We found that health centers with less reliable access to energy are staffed with predominantly female-dominated cadres such as nurses who are also less well-equipped to do their jobs based on the lack of consistent availability of equipment. Gender segregation in the health workforce also leads to worse working conditions, and having poor energy supply may impede women from doing their jobs [6]. Better electricity access can improve the retention of skilled health workers, extend night-time service provision, and allow for a faster emergency response including for childbirth emergencies [10]. Other similar female-dominated cadres such as midwives are more available at clinics with better electricity: in Haiti, the dispensaries with better electricity access had four times as many degree midwives on average. In the DRC, health centers with grid access have more nurses on average than those that do not.

Female-dominated professions are at the frontlines of providing primary care to women, infants, and children. Reduced availability of female medical staff at health centers with poor energy access therefore has important implications for maternal and health. If fewer facilities are capable of conducting normal deliveries, in part due to poor electricity access, this means women have to travel farther and farther from their villages for safe childbirth. In the DRC, 74% of the population lives more than 5 km from a health center and our research shows that if the closest health center does not have reliable access to energy, certain equipment central to providing high quality maternal care (e.g. ultrasound, delivery light) may not be available [45] Thus, the quality of care may be impacted by the availability of equipment and health workforce dynamics. A multi-country study focused on African nations founds that the quality of antenatal care is better at better-equipped and better-staffed facilities, and that nurses provide higher quality care [33]. This makes having safe working environments for cadres that are heavily female-dominated crucial, especially because of their role in the delivery of primary care services including MCH care.

In conclusion, our comparative analysis shows that depending on the level of energy access at primary level health facilities, there are observable disparities in health service delivery, medical equipment availability and health workforce capacity. We further show that these disparities disproportionately affect women, as both providers and seekers of primary healthcare for themselves and their children. Due to limitations with the available data, we are unable to estimate a causal relationship between electricity access and healthcare. However, our findings demonstrate that this relationship is important to understand and study further. Public health researchers should focus studying the causal relationship between energy and health system outcomes, identifying potential endogenous factors that may be explaining the relationship, and more in-depth gender analysis on the impacts of low/unreliable energy access especially on the health workforce. WHO and other practitioners recognize a lack of reliable energy access as a barrier to basic healthcare delivery globally, but more research is necessary to understand these linkages further as a precursor to the design effective solutions and policies. While strengthening the energy infrastructure is not sufficient to strengthen health systems alone, it is absolutely a necessary condition to building resilient health systems in LMICs, and decentralized sustainable energy systems provide a demonstrated pathway for catalyzing further improvements in the health systems. [5]. Better infrastructure at the lower levels, enabled by better energy access, can bring services delivered predominantly by women closer to the women who need them the most.

## Data Availability

All data used in this study are publicly available at the URL mentioned herein.

https://dhsprogram.com/data/available-datasets.cfm

## Acknowledgements

VS acknowledges support from the J.J. “Jake” Pickle Doctoral Fellowship. SMM acknowledges support from the grant P2CHD042849, Population Research Center, awarded to the Population Research Center at The University of Texas at Austin by the Eunice Kennedy Shriver National Institute of Child Health and Human Development. This research has also received support from the grant, T32HD007081, Training Program in Population Studies, awarded to the Population Research Center at The University of Texas at Austin by the Eunice Kennedy Shriver National Institute of Child Health and Human Development. The content is solely the responsibility of the authors and does not necessarily represent the official views of the National Institutes of Health.

## Footnotes

1 Cesarean sections are typically conducted only in referral hospitals and secondary facilities, so we would not expect any of the primary level facilities to provide this service. However, we included it in the analysis here since one in ten health centers in the DRC actually reported that they offer C-sections as well.

## References

1. World Health Organization. Primary health care [Internet]. 2020 [cited 2020 Jun 11]. Available from: https://www.who.int/westernpacific/health-topics/primary-health-care

2. Gage AD, Leslie HH, Bitton A, Jerome JG, Joseph JP, Thermidor R, et al. Does quality influence utilization of primary health care? Evidence from Haiti. Global Health [Internet]. 2018 Jun 20 [cited 2019 Sep 23];14. Available from: https://www.ncbi.nlm.nih.gov/pmc/articles/PMC6011404/

3. World Health Organization. Maternal health [Internet]. 2020 [cited 2020 Jul 9]. Available from: https://www.who.int/westernpacific/health-topics/maternal-health

4. Office of Disease Prevention and Human Promotion. Maternal, Infant, and Child Health [Internet]. Healthy People 2020. 2020 [cited 2020 Jul 9]. Available from: https://www.healthypeople.gov/2020/topics-objectives/topic/maternal-infant-and-child-health

5. Javadi D, Ssempebwa J, Isunju JB, Yevoo L, Amu A, Nabiwemba E, et al. Implementation research on sustainable electrification of rural primary care facilities in Ghana and Uganda. Health Policy and Planning. 2020;35(Supplement_2):ii124–36.

6. World Health Organization. Delivered by women, led by men: A gender and equity analysis of the global health and social workforce. 2019;

7. Morgan R, Ayiasi RM, Barman D, Buzuzi S, Ssemugabo C, Ezumah N, et al. Gendered health systems: evidence from low-and middle-income countries. Health research policy and systems. 2018;16(1):58.

8. Shastry V, Rai V. Reduced health services at under-electrified primary healthcare facilities: Evidence from India. PLOS ONE. 2021 Jun 4;16(6):e0252705.

9. World Health Organization. Monitoring the building blocks of health systems: a handbook of indicators and their measurement strategies. World Health Organization; 2010.

10. World Health Organization. Access to modern energy services for health facilities in resource-constrained settings: a review of status, significance, challenges and measurement. 2014;

11. Adair-Rohani H, Zukor K, Bonjour S, Wilburn S, Kuesel AC, Hebert R, et al. Limited electricity access in health facilities of sub-Saharan Africa: a systematic review of data on electricity access, sources, and reliability. Global Health: Science and Practice. 2013;1(2):249–61.

12. Conceição P. Human Development Report 2019: Beyond Income, Beyond Averages, Beyond Today: Inequalities in Human Development in the 21st Century. United Nations Development Programme; 2019.

13. The World Bank. GDP per capita (current US$) [Internet]. 2020 [cited 2020 Jul 1]. Available from: https://data.worldbank.org/indicator/NY.GDP.PCAP.CD

14. The World Bank. Population, total [Internet]. 2020 [cited 2020 Jul 17]. Available from: https://data.worldbank.org/indicator/SP.POP.TOTL

15. UNICEF. Country profiles Haiti l Demographics, Health & Infant Mortality [Internet]. UNICEF DATA. 2020 [cited 2020 Jul 9]. Available from: https://data.unicef.org/country/hti/

16. UNICEF. Country profiles Senegal Demographics, Health & Infant Mortality [Internet]. UNICEF DATA. 2020 [cited 2020 Jul 10]. Available from: https://data.unicef.org/country/sen/

17. United Nations Development Programme. Human Development Index (HDI) [Internet]. United Nations Development Programme Human Development Reports. 2020 [cited 2020 Jul 9]. Available from: http://hdr.undp.org/en/content/human-development-index-hdi

18. Dowell SF, Tappero JW, Frieden TR. Public health in Haiti—challenges and progress. New England journal of medicine. 2011;364(4):300–1.

19. WHO,. Haiti [Internet]. WHO. 2019 [cited 2019 Dec 16]. Available from: http://www.who.int/countries/hti/en/

20. The Centers for Disease Control and Prevention. Progress Towards Rebuilding Haiti’s Health System [Internet]. 2013. Available from: https://www.cdc.gov/globalhealth/healthprotection/errb/pdf/progresstowardrebuildinghaitis healthsystem.pdf

21. World Health Organization. Trends in maternal mortality 2000 to 2017: estimates by WHO, UNICEF, UNFPA, World Bank Group and the United Nations Population Division. 2019;

22. Institute for Health Metrics and Evaluation. Senegal [Internet]. Institute for Health Metrics and Evaluation. 2018 [cited 2020 Jul 10]. Available from: http://www.healthdata.org/senegal

23. The World Bank. Maternal mortality ratio (modeled estimate, per 100,000 live births) - Senegal [Internet]. 2019 [cited 2020 Jul 10]. Available from: https://data.worldbank.org/indicator/SH.STA.MMRT?locations=SN

24. World Health Organization. Senegal [Internet]. WHO. 2019 [cited 2019 Dec 16]. Available from: http://www.who.int/countries/sen/en/

25. Institute for Health Metrics and Evaluation. Democratic Republic of the Congo [Internet]. Institute for Health Metrics and Evaluation. 2018 [cited 2020 Jul 10]. Available from: http://www.healthdata.org/democratic-republic-congo

26. U.S. Agency for International Development. Health Fact Sheet: Democratic Republic of the Congo [Internet]. 2019 [cited 2020 Jun 26]. Available from: https://www.usaid.gov/democratic-republic-congo/fact-sheets/usaiddrc-fact-sheet-health

27. The Centers for Disease Control and Prevention. Democratic Republic of Congo - CDC Center for Global Health [Internet]. 2019 [cited 2019 Dec 16]. Available from: https://www.cdc.gov/globalhealth/countries/drc/default.htm

28. Pan American Health Organization. Haiti [Internet]. 2020 [cited 2020 Jun 26]. Available from: https://www.paho.org/salud-en-las-americas-2017/?p=4110

29. The Centers for Disease Control and Prevention. Remarkable Progress Five Years after Haiti Earthquake [Internet]. CDC. 2015 [cited 2020 Jul 10]. Available from: https://www.cdc.gov/media/releases/2015/p0112-haiti-earthquake.html

30. U.S. Agency for International Development. Haiti-Overview [Internet]. 2020 [cited 2020 Jul 10]. Available from: https://www.usaid.gov/where-we-work/latin-american-and-caribbean/haiti/haiti-overview

31. Azevedo MJ. Historical Perspectives on the State of Health and Health Systems in Africa, Volume I: The Pre-Colonial and Colonial Eras. Springer; 2017.

32. Heyen-Perschon J. Report on current situation in the health sector of Senegal and possible roles for non-motorised transport interventions. Institute for Transportation and Development Policy. 2005;

33. Kruk ME, Chukwuma A, Mbaruku G, Leslie HH. Variation in quality of primary-care services in Kenya, Malawi, Namibia, Rwanda, Senegal, Uganda and the United Republic of Tanzania. Bull World Health Organ. 2017 Jun 1;95(6):408–18.

34. Ntembwa HK, Lerberghe WV. Demogratic Republic of the Congo Improving aid coordination in the health sector [Internet]. World Health Organization,; 2015. (Improving Health System Efficiency). Available from: https://apps.who.int/iris/bitstream/handle/10665/186673/WHO_HIS_HGF_CaseStudy_15.4 _eng.pdf?sequence=1

35. Ho LS, Labrecque G, Batonon I, Salsi V, Ratnayake R. Effects of a community scorecard on improving the local health system in Eastern Democratic Republic of Congo: qualitative evidence using the most significant change technique. Conflict and Health. 2015 Sep 3;9(1):27.

36. Laokri S, Soelaeman R, Hotchkiss DR. Assessing out-of-pocket expenditures for primary health care: how responsive is the Democratic Republic of Congo health system to providing financial risk protection? BMC Health Services Research. 2018 Jun 15;18(1):451.

37. Kalisya LM, Salmon M, Manwa K, Muller MM, Diango K, Zaidi R, et al. The state of emergency care in Democratic Republic of Congo. African Journal of Emergency Medicine. 2015 Dec 1;5(4):153–8.

38. ICF. Demographic and Health Surveys (various) [Datasets]. Funded by USAID. Rockville, Maryland: ICF [Distributor].; 2018.

39. Julious SA. Using confidence intervals around individual means to assess statistical significance between two means. Pharmaceutical Statistics. 2004;3(3):217–22.

40. Leslie HH, Sun Z, Kruk ME. Association between infrastructure and observed quality of care in 4 healthcare services: A cross-sectional study of 4,300 facilities in 8 countries. PLoS medicine. 2017 Dec;14(12):e1002464.

41. Ivers LC. Strengthening the health system while investing in Haiti. Am J Public Health. 2011 Jun;101(6):970–1.

42. Jerome J, Ivers LC. Community health workers in health systems strengthening: a qualitative evaluation from rural Haiti. AIDS (London, England). 2010;24(Suppl 1):S67.

43. Bertone MP, Lurton G, Mutombo PB. Investigating the remuneration of health workers in the DR Congo: implications for the health workforce and the health system in a fragile setting. Health Policy and Planning. 2016 Jan 11;31(9):1143–51.

44. Zurn P, Codjia L, Sall FL, Braichet J-M. How to recruit and retain health workers in underserved areas: the Senegalese experience. Bulletin of the World Health Organization. 2010;88:386–9.

45. Ministere de la Sante Publique. Plan national de développement sanitaire (PNDS): 2011– 2015. Kinshasa: Ministere de la Sante Publique [République Démocratique du Congo] Secretariat General; 2010.

